# Comparison of SARS-CoV-2 detection in Saliva by real-time RT-PCR and RT-PCR/MALDI-TOF Methods

**DOI:** 10.1101/2021.03.11.21253234

**Authors:** Matthew M. Hernandez, Radhika Banu, Paras Shrestha, Armi Patel, Feng Chen, Liyong Cao, Shelcie Fabre, Jessica Tan, Heidi Lopez, Numthip Chiu, Biana Shifrin, Inessa Zapolskaya, Vanessa Flores, Pui Yiu Lee, Sergio Castañeda, Juan David Ramírez, Jeffrey Jhang, Giuliana Osorio, Melissa R. Gitman, Michael D. Nowak, David L. Reich, Carlos Cordon-Cardo, Emilia Mia Sordillo, Alberto E. Paniz-Mondolfi

## Abstract

The coronavirus disease 2019 (COVID-19) pandemic has accelerated the need for rapid implementation of diagnostic assays for detection of severe acute respiratory syndrome coronavirus-2 (SARS-CoV-2) in respiratory specimens. While multiple molecular methods utilize nasopharyngeal specimens, supply chain constraints and need for easier and safer specimen collection warrant alternative specimen types, particularly saliva. Although saliva has been found to be a comparable clinical matrix for detection of SARS-CoV-2, evaluations of diagnostic and analytic performance across platforms for this specimen type are limited. Here, we compared two methods for SARS-CoV-2 detection in saliva: the Roche cobas® 6800/8800 SARS-CoV-2 real-time RT-PCR Test and the Agena Biosciences MassARRAY^®^ SARS-CoV-2 Panel/MassARRAY^®^ System. Overall, both systems had high agreement with one another, and both demonstrated high diagnostic sensitivity and specificity when compared to matched patient upper respiratory specimens. We also evaluated the analytical sensitivity of each platform and determined the limit of detection of the Roche assay was four times lower than that of Agena for saliva specimens (390.6 v. 1,562.5 copies/mL). Furthermore, across individual target components of each assay, T2 and N2 targets had the lowest limits of detection for each platform, respectively. Together, we demonstrate that saliva represents an appropriate specimen for SARS-CoV-2 detection in two technologies that have high agreement and differ in analytical sensitivities overall and across individual component targets. The addition of saliva as an acceptable specimen and understanding the sensitivity for testing on these platforms can further inform public health measures for screening and detection to combat the COVID-19 pandemic.

## INTRODUCTION

Accurate and rapid testing is vital to informing the response to the coronavirus disease 2019 (COVID-19) pandemic. Since its inception, nucleic acid amplification testing (NAAT) for SARS-CoV-2 RNA in nasopharyngeal (NP) specimens has been the mainstay in diagnosing COVID-19. Collection of such specimens requires sampling by trained healthcare professionals who need materials such as swabs and viral transport medium (VTM) that may not be available in all settings (1–3). Currently, alternative specimen types including anterior nares (AN) and oropharyngeal (OP) specimens have been evaluated and approved for testing.

Saliva has recently garnered attention as a potential specimen given its lower discomfort, minimal invasiveness, and ability to be self-collected. As of February 21, 2021, nineteen *in vitro* SARS-CoV-2 diagnostic tests utilizing saliva as a clinical matrix have been approved for Emergency Use Authorization (EUA) by the U.S. Food and Drug Administration (4). Indeed, recent systematic reviews of reported studies demonstrated that saliva NAAT diagnostic performance is comparable to that of NP specimens, particularly in the ambulatory setting (5, 6). While studies have compared detection of SARS-CoV-2 across matched NP and saliva specimens, there is large variability in specimen collection, processing methods, and testing platforms utilized (5–14). Moreover, studies that assess analytical performance of detection in saliva across platforms are lacking.

Since the identification of SARS-CoV-2, high-throughput sample processing has been logistically difficult to achieve given a number of hurdles including instrument availability and supply chain limitations (15–18). Recently, a novel multiplex reverse transcription (RT-PCR)/MALDI-TOF assay from Agena Bioscience has received EUA (19). The MassARRAY^®^ SARS-CoV-2 Panel and MassARRAY^®^ System has the potential to increase diagnostic capacity and complement current standard NAAT technologies. This is particularly promising for use of saliva for large community-based testing efforts.

We therefore evaluated this platform (“Agena”) and the more ubiquitous cobas^®^ 6800/8800 SARS-CoV-2 real-time RT-PCR Test (“Roche”) to detect SARS-CoV-2 in saliva specimens. Furthermore, we also compared the analytic performance of each platform and each of its component targets.

## MATERIALS AND METHODS

We undertook a direct comparison of saliva as a clinical specimen for detection of SARS-CoV-2 viral nucleic acids across two platforms in the Clinical Microbiology Laboratory (CML) for the Mount Sinai Health System (MSHS) which is certified under Clinical Laboratory Improvement Amendments of 1988 (CLIA), 42 U.S.C. §263a and meets requirements to perform high-complexity tests.

### Saliva specimen collection and processing

Saliva specimens were collected from sixty patients who underwent molecular testing for SARS-CoV-2 in NP or AN specimens collected within the previous 48 hours. Saliva specimens were collected in sterile containers (Corning, 352070) and volumes ranged from 0.5 to 1.5 mL. Upon receipt in MSHS CML, 1 mL of viral transport media (VTM) (Hardy Diagnostics, R99) was added to each saliva specimen. These saliva-VTM specimens were vortexed for 30 seconds and 1 mL of each was incubated at 55°C for 15 minutes. Processed specimens subsequently underwent side-by-side SARS-CoV-2 nucleic acid detection across two different platforms.

### SARS-CoV-2 testing

For testing with the cobas^®^ 6800/8800 SARS-CoV-2 real-time RT-PCR Test (Roche, 09175431190), aliquots of processed saliva specimens were run as previously described for NP specimens (20). Briefly, the assay utilizes two targets to detect SARS-CoV-2 RNA: the SARS-CoV-2-specific Orf1ab gene (T1) and the pan-Sarbecovirus envelope E gene (T2). A result was deemed positive for SARS-CoV-2 if both T1 and T2 were detected, or if T1 was detected alone. A result was deemed presumptive positive if T2 was detected alone. A result was deemed negative if neither T1 nor T2 was detected. Target results were valid across all specimens run.

For testing with the MassARRAY^®^ SARS-CoV-2 Panel and MassARRAY^®^ System (Agena, CPM384), RNA was extracted from 300 μL of processed specimens using the chemagic™ Viral DNA/RNA 300 Kit H96 (PerkinElmer, CMG-1033-S) on the automated chemagic™ 360 instrument (PerkinElmer, 2024-0020), as per the manufacturer’s protocol. To serve as an internal control (IC), MS2 phage RNA was included in all extraction steps. Extracted RNA underwent reverse transcription PCR (RT-PCR) with iPLEX^®^ Pro chemistry to amplify the different Agena target regions per the manufacturer’s protocol. After the inactivation of unincorporated dNTPs by treatment with shrimp alkaline phosphatase (SAP), a sequence-specific primer extension step was performed, in which a mass-modified terminator nucleotide was added to the probe, using the supplied extension primers and iPLEX^®^ Pro reagents.

The extension products (analyte) were desalted, transferred to a SpectroCHIP^®^ Array (a silicon chip with pre-spotted matrix crystal) and then loaded into the MassARRAY^®^ Analyzer (a MALDI-TOF mass spectrometer). For the sample analysis, the analyte/matrix co-crystals were irradiated by a laser, inducing desorption and ionization. The positively charged molecules accelerated into a flight tube towards a detector. Separation occurred by time-of-flight, which is proportional to the mass of the individual molecules. After data processing, a spectral fingerprint was generated for each analyte that characterizes the mass/charge ratio of the molecules (x-axis) as well as their relative intensity (y-axis). Data acquired by the MassARRAY^®^ Analyzer was processed with the MassARRAY^®^ Typer software and then the SARS-CoV-2 Report software. The assay was designed to detect five viral targets: three in the nucleocapsid (N) gene (N1, N2, N3) and two in the Orf1ab gene (ORF1, Orf1ab). If the MS2 IC was detected, results were interpreted as positive if at least two targets were detected or negative if less than two targets were detected. If MS2 IC was not detected and no targets were detected, the result was interpreted as invalid and required rerunning of the specimen.

### Limit of detection of SARS-CoV-2 nucleic acid in saliva

The limit of detection (LoD) was determined across both platforms using known concentrations of a SARS-CoV-2 standard spiked into saliva clinical matrix.

Briefly, an in-house SARS-CoV-2 standard was generated by pooling 59 NP specimens that previously tested positive at MSHS CML (average T1 cycle threshold (Ct) = 17.53, average T2 Ct = 17.59). To quantitate the standard, three dilutions of the pooled sample were made (e.g., 1:50,000 (D1), 1:100,000 (D2), 1:200,000 (D3)) and run alongside serial dilutions of a commercially available standard (ZeptoMetrix, NATSARS(COV2)-ERC) on the Roche platform which has EUA from the FDA for SARS-CoV-2 detection in NP specimens. All reactions were run in triplicate and SARS-CoV-2-negative NP matrix served as the diluent. Concentrations of each standard dilution was determined by extrapolation from standard curves generated across T1 and T2 targets (Fig. S1) for each dilution. The stock concentration was, in turn, calculated as the average of the extrapolated stock concentrations determined at each dilution. Aliquots (50 μL) of this stock measurand were stored at −80°C to prevent multiple freeze-thaw cycles.

To simulate collection of saliva for testing, saliva from healthy donors was combined in equal parts with VTM and spiked with the SARS-CoV-2 measurand. Serial dilutions of the spiked saliva-VTM specimens were generated in 50-mL conical vials (Corning 352070) over a range of 3,125.0 – 97.7 copies/mL (cp/mL) and 12,500 – 195.3 cp/mL for testing on the Roche and Agena platforms, respectively. For each platform, ten replicates of each dilution were generated as well as ten replicates of saliva-VTM spiked with the SARS-CoV-2-negative NP diluent to serve as negative controls. Spiked saliva-VTM specimens were processed and run as described above. SARS-CoV-2 was not detected in any of the negative controls and all results were valid across both platforms.

For each platform, the LoD of each overall assay and each target were determined. The experimental LoD represents the lowest concentration with 95% detection. The probit LoD (and 95% fiduciary confidence intervals) was determined by 95% detection based on a probit regression model.

### Statistical analyses

For comparison of outcomes across both platforms, percent agreement and Cohen’s kappa (κ) statistic were calculated using the attribute agreement analysis on Minitab Statistical Software (19.2020.2.0). Normality was assessed by D’Agostino and Pearson test for continuous variables (e.g., Ct values) (GraphPad Prism 9.0.2). Student’s t-test (two-tailed) was performed if data was normally distributed; otherwise, the Mann-Whitney test (two-tailed) was utilized (GraphPad Prism 9.0.2). Simple linear regression analyses were performed across Roche Ct values and serial dilutions. Probit regression modeling assuming Weibull distribution was performed if at least two probit points were available (e.g., not 100% or 0% detection) (Minitab Statistical Software, 19.2020.2.0). Where depicted, confidence intervals (CI) reflect the 95% level.

## RESULTS

Sixty patients who underwent testing for SARS-CoV-2 by NAAT (NP or AN) at MSHS CML were provided with sterile containers for submission of saliva specimens within 48 hours of diagnosis. Saliva specimens were immediately processed and run side-by-side on the Roche and Agena platforms. When compared to paired NP or AN specimens, both platforms had equivalent sensitivities (97.14%, CI: 85.08-99.93%) and specificities (100%, CI: 86.28-100%) for saliva specimens.

The Roche platform detected SARS-CoV-2 RNA in 34/60 saliva specimens (Table 1). Of the remaining 26, two specimens resulted as presumptive positive and were considered not detected for this study. The Agena platform detected SARS-CoV-2 RNA in 34/60 specimens. Of note, one of the two presumptive positive specimens by Roche was detected by Agena. In addition, the one specimen detected by Roche but not by Agena had the highest T1 Ct (31.62) and second highest T2 Ct (33.68) of all specimens tested. Overall, there was an almost perfect level of agreement across the two platforms (96.67% agreement, CI: 88.47-99.59; Cohen’s κ = 0.9321, p = 2.6×10^−13^).

**Table 1.** Detection of SARS-CoV-2 nucleic acids in saliva across Roche and Agena commercial systems.

|  |  | **Roche** |  |  |
| --- | --- | --- | --- | --- |
|  |  | Positive | Negative | **Total** |
| **Agena** | Positive | 33 | 1 * | 34 |
|  | Negative | 1 | 25 | 26 |
|  | **Total** | 34 | 26 | 60 |

\*Presumptive positive by Roche

To preliminarily assess the sensitivity of each platform, we evaluated the performance of component targets across the saliva clinical specimens (Fig. 1). Roche Ct values for each target ranged from 18.80-31.62 for target T1 (Orf1ab gene) and 19.06-37.46 for target T2 (E gene). When compared to the number of targets detected on the Agena platform, all five Agena targets were detected in specimens that had the lowest mean (±SD) Ct values on Roche T1 (24.64±3.019) and T2 (25.26±3.189) targets. The number of Agena targets detected in clinical saliva specimens progressively decreased with increasing Ct values across both Roche targets.

**Fig 1.**
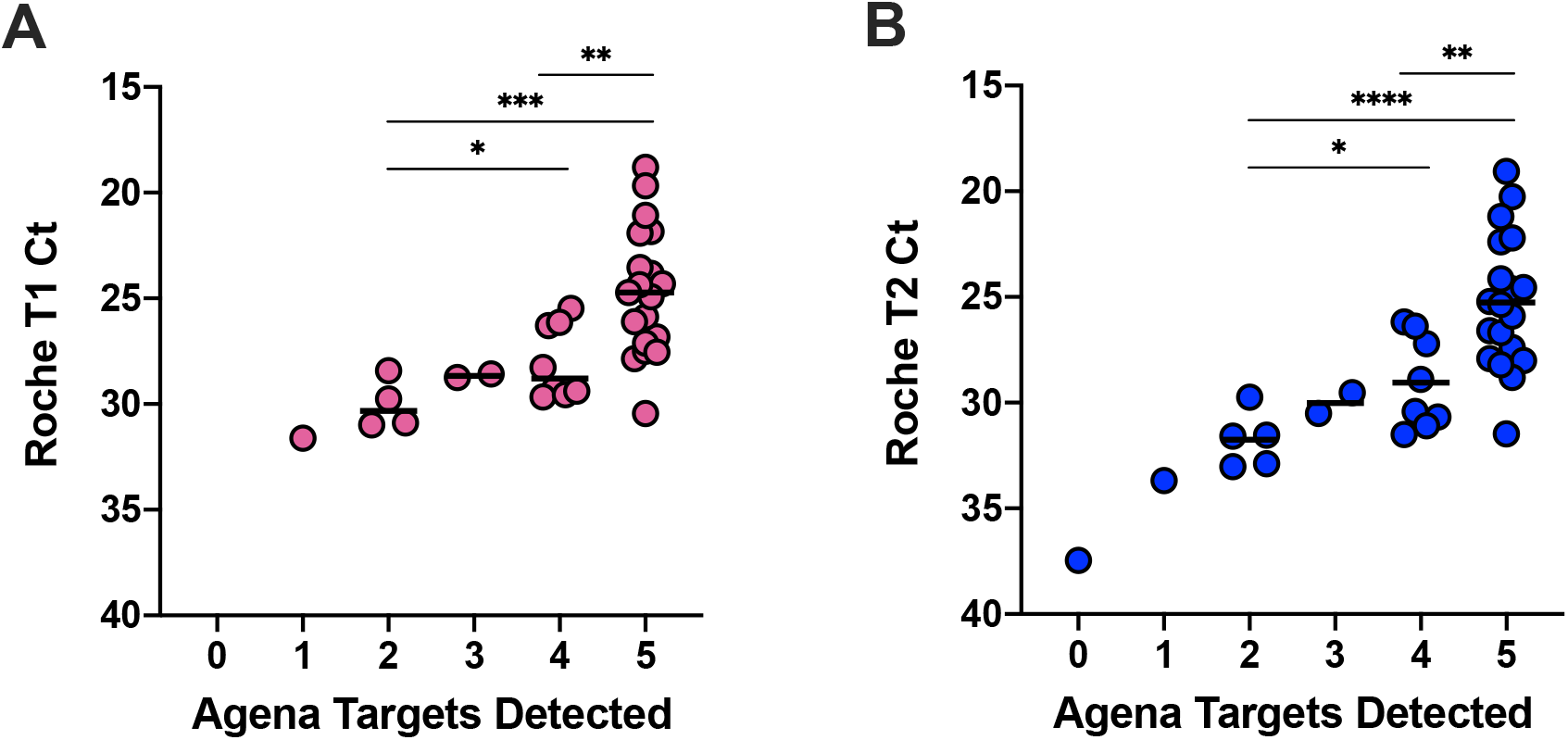
Quantitative comparison of SARS-CoV-2 targets detected in clinical saliva specimens. Scatter plots depict the number of SARS-CoV-2 targets on the Agena platform detected and the corresponding Roche cycle threshold (Ct) for each clinical saliva specimen. (A) Ct values for Roche target T1 (Orf1ab) and (B) Roche target T2 (E gene) are depicted for individual clinical saliva specimens. Medians are depicted in each column. Statistically significant differences are depicted (e.g., *, p<0.05; **, p<0.01; ***, p<0.001, ****, p<0.0001) based on student’s t-test or Mann-Whitney non-parametric test depending on whether data was normally distributed (see Methods).

We next systematically measured the limit of detection (LoD) of each platform and the component targets. We generated a SARS-CoV-2 standard from high-titer positive NP specimens collected from MSHS patients diagnosed at CML. The titer of the in-house standard was determined by extrapolating concentrations of three dilutions run alongside serial dilutions of a commercial SARS-CoV-2 standard on the Roche platform (Fig. S1). This had the benefit of accounting for any variation in extraction efficiency. The in-house standard was spiked into saliva matrix collected from healthy donors and ten replicates of serial dilutions were run side-by-side on each platform. On the Agena platform, the experimental LoD was determined to be 1,562.5 cp/mL (Table 2) which is slightly lower than the LoD reported by manufacturers for NP clinical matrix (2,500 cp/mL) (19). Across the five different Agena targets, the most sensitive target was the N2 target (1,562.5 cp/mL) followed by the N1 target (3,125 cp/mL) (Table 2, Fig. 2A). The least sensitive was the Orf1ab target whose LoD could not be determined from the range of concentrations tested. This reflected a gradient in performance across the individual components on the Agena platform.

**Table 2.**
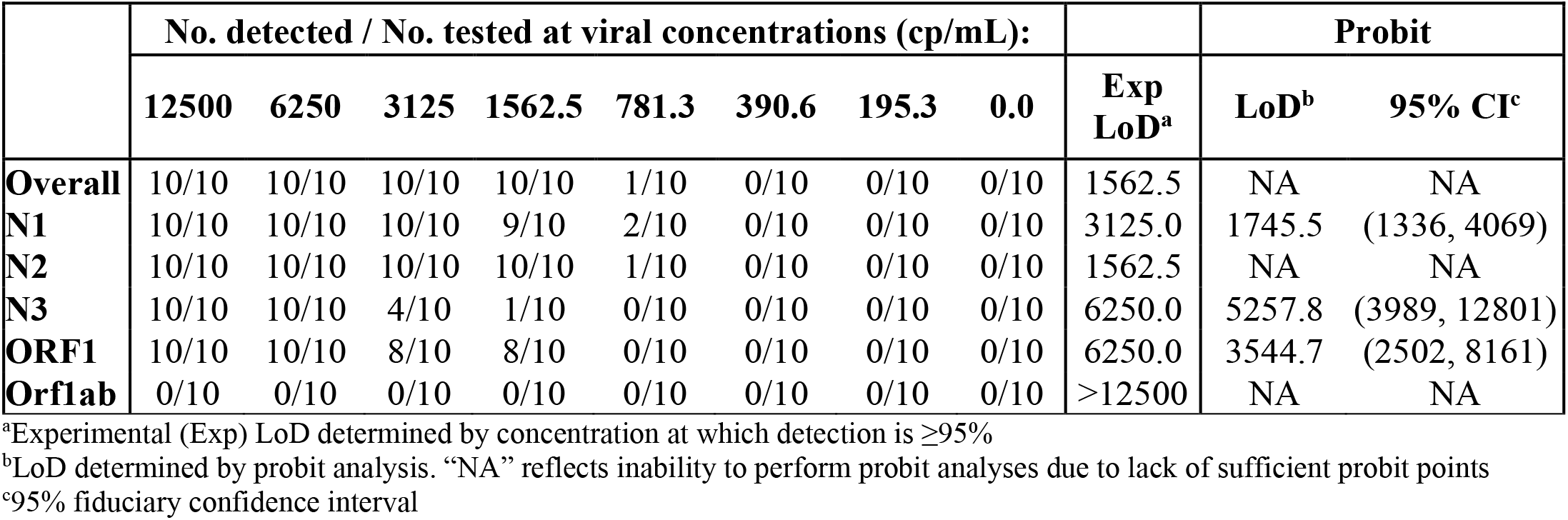
LoD of SARS-CoV-2 nucleic acids in spiked saliva on the Agena MassARRAY^®^ platform.

**Fig 2.**
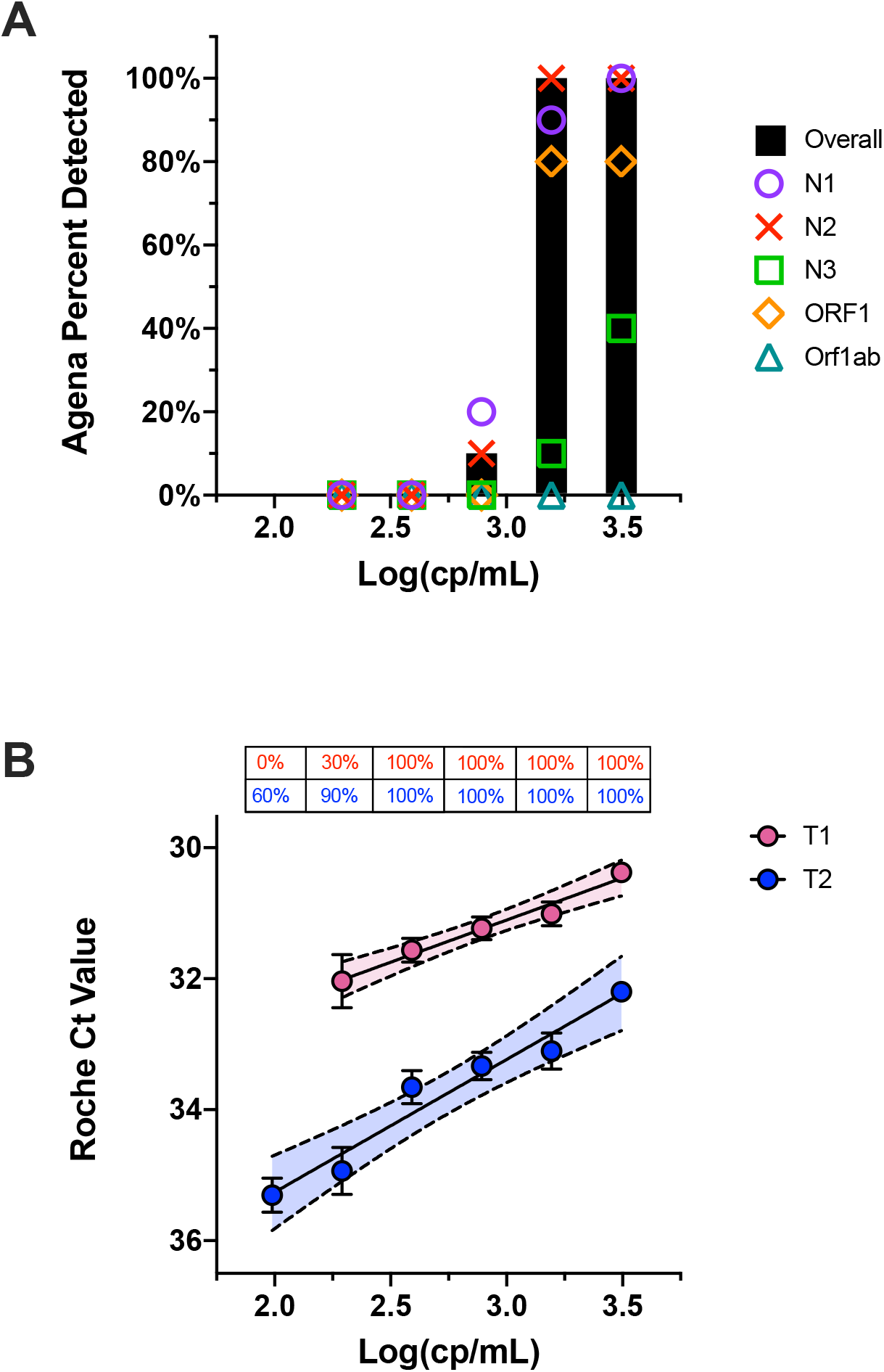
Evaluation of Roche and Agena SARS-CoV-2 target sensitivity. (A) Bar graph depicts percent of spiked saliva specimens detected overall by the Agena MassARRAY^®^ platform at five different concentration (log). Overlaid are the individual sensitivities of the five Agena targets at each concentration. (B) Scatter plot of Ct values of Roche T1 (pink) and T2 (blue) targets across concentrations (log) of spiked saliva specimens at six different concentrations. Mean, standard error of the mean, and line of best fit with 95% confidence intervals are depicted for each target. Above each concentration is the percent of replicates detected by T1 or T2 targets.

On the Roche platform, the experimental LoD was lower than that of Agena at 390.6 cp/mL. The Ct values for these saliva specimens demonstrated a linear correlation with the corresponding concentrations across both T1 (R^2^=0.9760, p=0.0016) and T2 (R^2^=0.9534, p=0.008) (Fig. 2B). Overall, T2 Ct values were higher than T1 Ct values for specimens at the same concentration (p<0.01) which is consistent with previous reports for NP specimens (21, 22). While the experimental LoD for T1 and T2 targets were determined equivalent, probit analyses suggest the LoD of T2 is, in fact, lower (228.6 cp/mL). However, the fiduciary confidence interval for this value is broad (151.4-3.7×10^10^) given that the concentration at which no specimens were detected was not determined in our study.

## DISCUSSION

Saliva represents an attractive alternative specimen type for SARS-CoV-2 testing given its limited invasiveness, ability to be self-collected, and reduced need for limited supplies A number of groups have demonstrated that saliva is an acceptable and sensitive specimen type when compared to other upper respiratory (e.g., NP, AN, OP) specimens (5, 8, 9, 12–14). However, analytical performance of this specimen type has yet to be evaluated across the multitude of platforms utilized. In this study, we demonstrate the utility of saliva as a diagnostic specimen across the Roche and Agena platforms. Saliva specimens collected within two days are equivocally sensitive and specific across both methods when compared to matched NP or AN specimens.

It is important to note that these two platforms tested are distinguished from each other by their technologic basis and their molecular targets. The Roche platform is like most of the current SARS-CoV-2 molecular diagnostic assays in that it utilizes real-time RT-PCR for detection. However, the Agena platform utilizes mass spectrometry to detect targeted amplicons produced by RT-PCR. While distinct in platform technology, our findings demonstrate comparable diagnostic capabilities of both platforms for detection of SARS-CoV-2 nucleic acids in clinical saliva specimens (Table 1).

The platforms we evaluated also differed by SARS-CoV-2 viral targets probed. In contrast to the Roche platform, which is based on two target amplicons (SARS-CoV-2 Orf1ab (T1) and pan-Sarbecovirus E genes (T2)), the Agena platform probes for five targets across two viral genes (3 targets in the nucleocapsid gene (N1, N2, N3), 2 targets in the Orf1ab gene (ORF1, Orf1ab)). This redundancy in viral targets is required to ensure robust sensitivity. When we assessed analytic performance of each target across the clinical saliva specimens, we observed variation in target performance with decreasing viral titers (e.g., Ct values), particularly within the Agena platform (Fig. 1). Specifically, the number of Agena targets detected progressively dropped with decreasing concentration. This suggested inherent analytic differences in the component targets that warrant further investigating.

In order to effectively utilize saliva as a clinical specimen for SARS-CoV-2 testing, it is essential to characterize the analytical sensitivity for each diagnostic platform. Most studies have yet to evaluate the LoD across platforms in a standardized method for saliva specimens (reviewed in (6)). Moreover, analytic sensitivity of component targets are often reported as those described by manufacturers or are not systematically evaluated, reported, nor compared across platforms (6, 7, 23–25). Our study demonstrates a greater sensitivity in the Roche platform for saliva specimens overall (Fig. 2). We also demonstrate that across both platforms, there are some targets which are more sensitive than others such as N2 in Agena and T2 in Roche (Fig. 2, Table 2). These metrics are vital as they can inform how diagnostic labs address new circulating viral variants that have mutations that may interfere with multiple detection methods.

Our study does have limitations in that our saliva collection methods did not occur at one time point but rather at any point in the day within two days of initial NP/AN collection. While the utility of standardized collection methods (e.g., early morning collection) remain to be further clarified, this is not a variable we controlled in this study. In addition, we utilized a pooled positive NP specimen to serve as our analyte to assess sensitivity. As a result, the sensitivities measured are based on a potentially heterogenous mixture of viral variants. We addressed this by pooling specimens isolated from two consecutive days to ensure a sampling of the predominant circulating clade virus at the given time period. Overall, we demonstrate comparable analytical performance across two unique diagnostic platforms for detection of SARS-CoV-2 nucleic acids in saliva specimens. Given the continued spread and rise of new SARS-CoV-2 variants, there is a critical need to understand the analytic capabilities of these technologies. This is especially relevant in large-scale screening efforts where saliva has the potential to be further exploited for its utility as a clinical specimen. This greater understanding of assay and target sensitivity is essential to informing both effective detection efforts and broader public health measures to ultimately quell the COVID-19 pandemic.

## Data Availability

The manuscript contains all relevant data.

## ACKNOWLEDGMENTS

We thank the members of the MSHS CML for providing any assistance when needed throughout this study. We also would like to thank the patients and healthy donors for providing specimens to complete this study.

## AUTHOR CONTRIBUTIONS

M.M.H., R.B., P.S., S.F., J.T., A.E.PM., H.L.,M.R.G., M.D.N., and E.M.S. provided clinical samples for the study. M.M.H., R.B., P.S., A.P., F.C., L.C., H.L., N.C., G.O., B.S., I.Z., V.F., P.Y., and, A.E.PM. accessioned clinical samples. M.M.H., R.B., P.S., A.P., F.C., L.C., and A.E.PM. performed limit of detection studies. M.M.H., L.S.G., J.D.R., J.J., D.L.R., C.C.C., E.M.S., and A.E.PM. analyzed, interpreted, or discussed data. M.M.H. and A.E.PM. wrote the manuscript. M.M.H., R.B., and A.E.PM. conceived the study. M.M.H., R.B., and A.E.PM. supervised the study. DLR raised financial support.

## SUPPLEMENTAL FIGURES

**Fig. S1.**
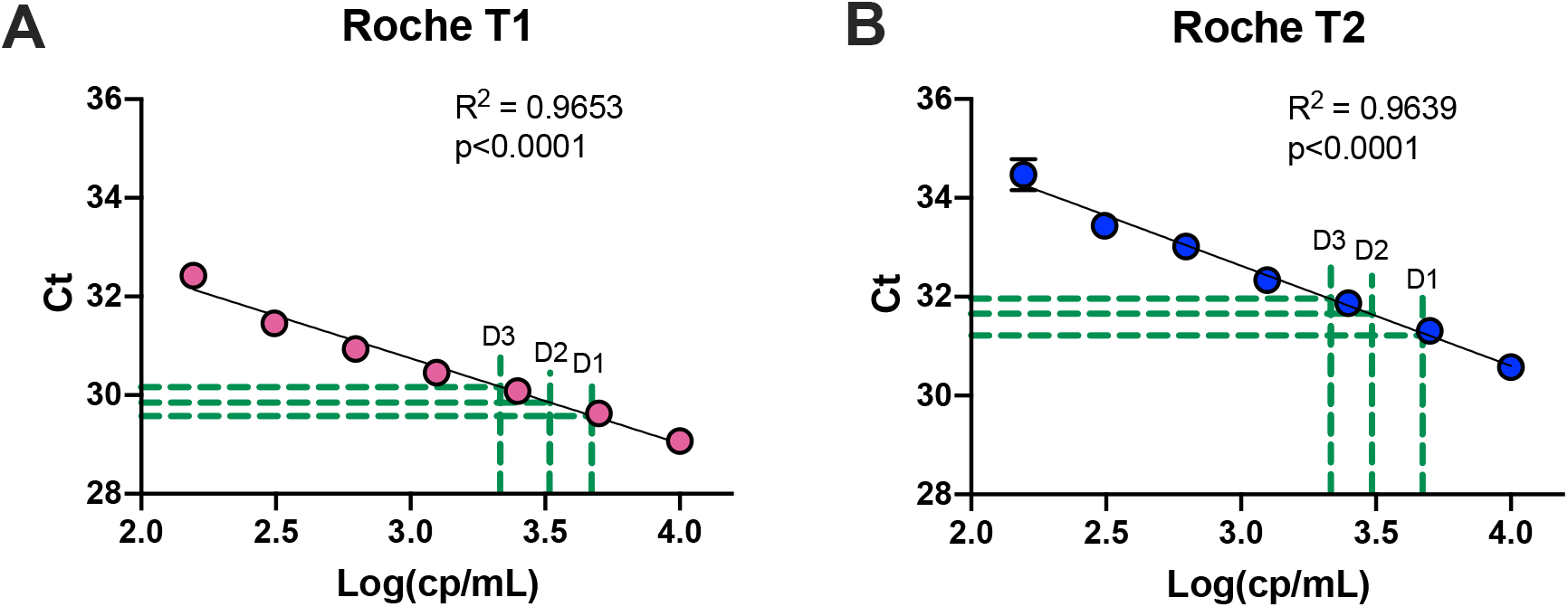
Quantitation of in-house SARS-CoV-2 standard. Linear regression of commercial standard in negative NP matrix run alongside three dilutions of pooled positive SARS-CoV-2 NP clinical specimens. (A) Mean (±SEM) Roche target T1 Ct values and (B) target T2 Ct values of seven serial dilutions of commercial standard plotted with lines of best fit. Correlation coefficients and p-values are annotated for each standard curve. Concentrations for three dilutions (D1, D2, D3) of in-house standard were extrapolated from T1 and T2 lines of best fit (dotted green lines) to determine the concentration of the in-house standard.

## REFERENCES

1. Lieberman JA, Pepper G, Naccache SN, Huang M-L, Jerome KR, Greninger AL. 2020. Comparison of Commercially Available and Laboratory-Developed Assays for In Vitro Detection of SARS-CoV-2 in Clinical Laboratories. J Clin Microbiol 58.

2. Kaul KL. 2020. Laboratories and Pandemic Preparedness: A Framework for Collaboration and Oversight. J Mol Diagn 22:841–843.

3. Zehnbauer B. 2021. Diagnostics in the Time of Coronavirus Disease 2019 (COVID-19): Challenges and Opportunities. J Mol Diagn 23:1–2.

4. Febraury 17, 2021. In Vitro Diagnostics EUAs. US Food and Drug Administration.

5. Butler-Laporte G, Lawandi A, Schiller I, Yao MC, Dendukuri N, McDonald EG, Lee TC. 2021. Comparison of Saliva and Nasopharyngeal Swab Nucleic Acid Amplification Testing for Detection of SARS-CoV-2: A Systematic Review and Meta-analysis. JAMA Intern Med https://doi.org/10.1001/jamainternmed.2020.8876.

6. Lee RA, Herigon JC, Benedetti A, Pollock NR, Denkinger CM. 2021. Performance of Saliva, Oropharyngeal Swabs, and Nasal Swabs for SARS-CoV-2 Molecular Detection: A Systematic Review and Meta-analysis. J Clin Microbiol https://doi.org/10.1128/JCM.02881-20.

7. Yee R, Truong TT, Pannaraj PS, Eubanks N, Gai E, Jumarang J, Turner L, Peralta A, Lee Y, Dien Bard J. 2021. Saliva Is a Promising Alternative Specimen for the Detection of SARS-CoV-2 in Children and Adults. J Clin Microbiol 59.

8. Procop GW, Shrestha NK, Vogel S, Van Sickle K, Harrington S, Rhoads DD, Rubin BP, Terpeluk P. 2020. A Direct Comparison of Enhanced Saliva to Nasopharyngeal Swab for the Detection of SARS-CoV-2 in Symptomatic Patients. J Clin Microbiol 58.

9. Wyllie AL, Fournier J, Casanovas-Massana A, Campbell M, Tokuyama M, Vijayakumar P, Warren JL, Geng B, Muenker MC, Moore AJ, Vogels CBF, Petrone ME, Ott IM, Lu P, Venkataraman A, Lu-Culligan A, Klein J, Earnest R, Simonov M, Datta R, Handoko R, Naushad N, Sewanan LR, Valdez J, White EB, Lapidus S, Kalinich CC, Jiang X, Kim DJ, Kudo E, Linehan M, Mao T, Moriyama M, Oh JE, Park A, Silva J, Song E, Takahashi T, Taura M, Weizman O-E, Wong P, Yang Y, Bermejo S, Odio CD, Omer SB, Dela Cruz CS, Farhadian S, Martinello RA, Iwasaki A, Grubaugh ND, Ko AI. 2020. Saliva or Nasopharyngeal Swab Specimens for Detection of SARS-CoV-2. N Engl J Med 383:1283–1286.

10. Ceron JJ, Lamy E, Martinez-Subiela S, Lopez-Jornet P, Capela E Silva F, Eckersall PD, Tvarijonaviciute A. 2020. Use of Saliva for Diagnosis and Monitoring the SARS-CoV-2: A General Perspective. J Clin Med Res 9.

11. Goldfarb DM, Tilley P, Al-Rawahi GN, Srigley JA, Ford G, Pedersen H, Pabbi A, Hannam-Clark S, Charles M, Dittrick M, Gadkar VJ, Pernica JM, Hoang LMN. 2021. Self-collected Saline Gargle Samples as an Alternative to Healthcare Worker Collected Nasopharyngeal Swabs for COVID-19 Diagnosis in Outpatients. J Clin Microbiol https://doi.org/10.1128/JCM.02427-20.

12. Zhu J, Guo J, Xu Y, Chen X. 2020. Viral dynamics of SARS-CoV-2 in saliva from infected patients. J Infect.

13. To KK-W, Tsang OT-Y, Leung W-S, Tam AR, Wu T-C, Lung DC, Yip CC-Y, Cai J-P, Chan JM-C, Chik TS-H, Lau DP-L, Choi CY-C, Chen L-L, Chan W-M, Chan K-H, Ip JD, Ng AC-K, Poon RW-S, Luo C-T, Cheng VC-C, Chan JF-W, Hung IF-N, Chen Z, Chen H, Yuen K-Y. 2020. Temporal profiles of viral load in posterior oropharyngeal saliva samples and serum antibody responses during infection by SARS-CoV-2: an observational cohort study. Lancet Infect Dis 20:565–574.

14. Teo AKJ, Choudhury Y, Tan IB, Cher CY, Chew SH, Wan ZY, Cheng LTE, Oon LLE, Tan MH, Chan KS, Hsu LY. 2021. Saliva is more sensitive than nasopharyngeal or nasal swabs for diagnosis of asymptomatic and mild COVID-19 infection. Sci Rep 11:3134.

15. Vandenberg O, Martiny D, Rochas O, van Belkum A, Kozlakidis Z. 2021. Considerations for diagnostic COVID-19 tests. Nat Rev Microbiol 19:171–183.

16. Lamprou DA. 2020. Emerging technologies for diagnostics and drug delivery in the fight against COVID-19 and other pandemics. Expert Rev Med Devices 17:1007–1012.

17. Younes N, Al-Sadeq DW, Al-Jighefee H, Younes S, Al-Jamal O, Daas HI, Yassine HM, Nasrallah GK. 2020. Challenges in Laboratory Diagnosis of the Novel Coronavirus SARS-CoV-2. Viruses 12.

18. Sheridan C. 2020. Coronavirus and the race to distribute reliable diagnostics. Nat Biotechnol 38:382–384.

19. Agena Bioscience, Inc. 2021. MassARRAY® SARS-CoV-2 Panel Instructions for Use.

20. Hernandez MM, Gonzalez-Reiche AS, Alshammary H, Fabre S, Khan Z, van De Guchte A, Obla A, Ellis E, Sullivan MJ, Tan J, Alburquerque B, Soto J, Wang C-Y, Sridhar SH, Wang Y-C, Smith M, Sebra R, Paniz-Mondolfi AE, Gitman MR, Nowak MD, Cordon-Cardo C, Luksza M, Krammer F, van Bakel H, Simon V, Sordillo EM. 2021. Before the surge: Molecular evidence of SARS-CoV-2 in New York city prior to the first report. bioRxiv. medRxiv.

21. Mostafa HH, Hardick J, Morehead E, Miller J-A, Gaydos CA, Manabe YC. 2020. Comparison of the analytical sensitivity of seven commonly used commercial SARS-CoV-2 automated molecular assays. J Clin Virol 130:104578.

22. Nalla AK, Casto AM, Huang M-LW, Perchetti GA, Sampoleo R, Shrestha L, Wei Y, Zhu H, Jerome KR, Greninger AL. 2020. Comparative Performance of SARS-CoV-2 Detection Assays Using Seven Different Primer-Probe Sets and One Assay Kit. J Clin Microbiol 58.

23. SoRelle JA, Mahimainathan L, McCormick-Baw C, Cavuoti D, Lee F, Bararia A, Thomas A, Sarode R, Clark AE, Muthukumar A. 2020. Evaluation of symptomatic patient saliva as a sample type for the Abbott ID NOW COVID-19 assay. bioRxiv. medRxiv.

24. Lu J, Becker D, Sandoval E, Amin A, De Hoff P, Diets A, Leonetti N, Lim YW, Elliott C, Laurent L, Grzymski J. 2020. Saliva is less sensitive than nasopharyngeal swabs for COVID-19 detection in the community setting. bioRxiv. medRxiv.

25. Pasomsub E, Watcharananan SP, Boonyawat K, Janchompoo P, Wongtabtim G, Suksuwan W, Sungkanuparph S, Phuphuakrat A. 2021. Saliva sample as a non-invasive specimen for the diagnosis of coronavirus disease 2019: a cross-sectional study. Clin Microbiol Infect 27:285.e1-285.e4.

